# Mood symptoms and chronic fatigue syndrome due to relapsing remitting multiple sclerosis are associated with immune activation and aberrations in the erythron

**DOI:** 10.1101/2022.12.30.22284056

**Authors:** Abbas F. Almulla, Al-Karrar Kais Abdul Jaleel, Ali Abbas Abo Algon, Chavit Tunvirachaisakul, Hayder K. Hassoun, Hussein K. Al-Hakeim, Michael Maes

## Abstract

**Background:** Multiple sclerosis (MS) is a chronic autoimmune and neuroinflammatory disease of the central nervous system characterized by peripheral activation of immune-inflammatory pathways which culminate in neurotoxicity causing demyelination of central neurons. Nonetheless, the pathophysiology of relapsing-remitting MS (RRMS)-related chronic fatigue, depression, anxiety, cognitive impairments, and autonomic disturbances is not well understood.

**Objectives:** The current study aims to delineate whether the remitted phase of RRMS is accompanied by activated immune-inflammatory pathways and if the latter, coupled with erythron variables, explain the chronic fatigue and mood symptoms due to RRMS.

**Material and Methods:** We recruited 63 MS patients, 55 in the remitted phase of RRMS and 8 with secondary progressive MS, and 30 healthy controls and assessed erythron variables and used a bio-plex assay to measure 27 serum cytokines.

**Results:** A significant part of the MS patients (46%) displayed activation of the immune-inflammatory response (IRS) and compensatory immune response (CIRS) systems, T helper (Th)1 and Th-17 cytokine profiles. Remitted RRMS patients showed increased chronic fatigue, depression, anxiety, physiosomatic, autonomic, and insomnia scores, which could partly be explained by M1 macrophage, Th1, Th-17, growth factor, and CIRS activation, as well as aberrations in the erythron including lowered hematocrit and hemoglobin levels.

**Conclusions:** Around 50% of remitted RRMS patients show activation of immune-inflammatory pathways in association with mood and chronic-fatigue-like symptoms. IRS and CIRS activation as well as the aberrations in the erythron are new drug targets to treat chronic fatigue and affective symptoms due to MS.

## Introduction

Multiple sclerosis (MS) is a chronic disease characterized by a cluster of symptoms comprising ataxia, numbness, visual disturbances, loss of balance, unexplained pain, chronic fatigue, muscle spasms, bladder malfunction, depression, cognitive impairments and progressing disabilities (Havrdová and Preiningerova, 2013; Jongen et al., 2012; Kallaur et al., 2016). MS shows a higher prevalence in high-income populations (North America 140/100.000 and Europe: 108/100.000) compared with 2.2 and 2.1/100,000 in East Asia and sub-Saharan Africa, respectively (Leray et al., 2016). In Iraq, a recent study revealed 4355 MS cases with a female/male ratio of 2.18:1 (Hassoun et al., 2022).

Based on the pattern of the disease, MS can be classified into relapsing-remitting MS (RRMS), secondary progressive MS (SPMS), primary progressive MS (PPMS), and clinically isolated syndromes (CIS) (Lublin and Reingold, 1996). While RRMS is the most prevalent MS phenotype, some of these patients develop SPMS accompanied by progressive deterioration leading to loss of neurological functions (Yiangou et al., 2006). Two mechanisms explain MS-associated disabilities as evaluated by extended disability scale scores (EDSS) (Kurtzke, J. F., 1983), namely gradual accumulation of dysfunctions resulting from poor recovery from acute relapses (denoted as relapse-associated worsening, RAW) and progression independent of relapse activity (PIRA) (Lublin et al., 2022; Roxburgh et al., 2005). MS disabilities are cumulative and the speed of their accumulation can be assessed by the multiple sclerosis severity scale (MSSS) (Roxburgh et al., 2005).

MS is a chronic autoimmune and neuroinflammatory disease of the central nervous system (CNS) and spinal cord resulting in demyelination of neurons (de Carvalho Jennings Pereira, Wildéa Lice et al., 2020; Siotto et al., 2019). Autoimmunity, neuroinflammation, oxidative stress damage, excitotoxicity, axonal and neuronal damage, demyelination, neurodegeneration, glial scar formation, remyelination, impairments of metabolic and mitochondrial metabolism are strongly involved in the acute relapses and progression of MS (Abbasi et al., 2016; Debouverie, 2009) (Morris, G. and Maes, M., 2013; Sospedra and Martin, 2016). Increased levels of thirteen cerebrospinal fluid (CSF) and twenty-one blood cytokines were reported in people with MS (Bai et al., 2019). For example, tumor necrosis factor (TNF)-α, CXCL-8, interleukin (IL)-15, IL-12p40, and CXCL13 were found to be elevated in CSF and blood (Bai et al., 2019).

A high burden of chronic fatigue, depression and anxiety is reported in MS and may impair the patient’s quality of life and interfere with the remission potential and thus prognosis of the illness (Boeschoten et al., 2017; Morris, G. et al., 2018; Nagaraj et al., 2013; Ormstad, H. et al., 2020). The prevalence of chronic fatigue, depression and anxiety in MS patients is between 75-90%, 27.01% and 35.19%, respectively (Ayache et al., 2022; Peres et al., 2022). Depression and anxiety may be more prevalent in PMS than in RRMS (Peres, Rodrigues et al. 2022), while there are no differences in chronic fatigue between both MS subtypes (Herring et al., 2021). Activated immune-inflammatory and oxidative and nitrosative stress (O&NS) pathways may partially explain the onset of mood symptoms and chronic fatigue in MS (Kallaur et al., 2016; Morris, G. et al., 2018; Ormstad, Heidi et al., 2020). Elevated concentrations of M1 macrophage and T helper (Th)1 cytokines along with high levels of reactive oxygen species (ROS), lipid peroxidation and nitric oxide (NO) play a role in depression due to MS (Kallaur et al., 2016). Moreover, a significant correlation was found between the severity of depression due to MS and increased expression of TNF-α and IFN-γ genes (Kahl et al., 2002; Pokryszko-Dragan et al., 2012). Plenty of studies indicate that chronic fatigue syndrome, major depressive disorder (MDD), and generalized anxiety disorder are associated with activated immune-inflammatory and O&NS pathways (Maes and Carvalho, 2018; Maes et al., 1998; Michopoulos et al., 2017; Vasupanrajit et al., 2022). For example, clinical depression is accompanied by increased levels of interleukin (IL)-1β, IL-6, IL-8, IL-17, IFN-γ, and TNF-α (Maes and Carvalho, 2018).

In general, it is more difficult to interpret solitary cytokine levels and, therefore, it is more adequate to describe cytokine findings in terms of profiles, including M1 macrophage, Th1, Th2, Th17, T regulatory (Treg), growth factor, and T cell growth factor profiles and to combine these further into composites reflecting activation of the immune-inflammatory response system (IRS) versus the compensatory immunoregulatory system (CIRS) (Maes and Carvalho, 2018). The latter system comprises immunoregulatory and anti-inflammatory mechanisms such as Th2 and Treg cytokines (e.g. IL-4 and IL-10) or soluble receptors (e.g. soluble IL-1 receptor antagonist (sIL-1RA) and sIL-2R), which tend to downregulate the IRS and counteract an overzealous inflammatory response (Maes and Carvalho, 2018). In this respect, MS is associated with activation of Th1, Th17, and Th1-like Th17, Th9, and Th22 profiles, whilst also Tregs play a key role (Kunkl et al., 2020) (Maciak et al., 2021; Melnikov, Mikhail and Lopatina, Anna, 2022). Clinical depression is accompanied by activated M1, Th1, Th17, T cell growth, IRS and CIRS profiles, although during acute episodes the IRS/CIRS ratio is increased (Maes and Carvalho, 2018). Nevertheless, there are no data on these profiles in depression, anxiety, and chronic fatigue due to RRMS.

Aberrations in the erythron may be involved in the pathophysiology of MS and depression. For example, Hon et al. found low levels of hemoglobin (Hb) in MS patients compared with healthy controls along with a significant inverse correlation between red blood cells (RBC) count and EDSS scores (Hon et al., 2012). Moreover, the RBC count is significantly decreased in patients with RRMS as compared to other types of MS and healthy controls (Kasprzycka et al., 2019). Depressed patients have significantly lowered RBCs, hematocrit (Hct) and Hb levels compared with healthy controls, whereas RDW and reticulocytes are significantly elevated, suggesting inflammation-associated anemia (Maes, M. et al., 1996; Vandoolaeghe et al., 1999; Wysokiński and Szczepocka, 2018). However, there are no data on the association between erythron features and IRS activation, chronic fatigue, and mood symptoms due to RRMS.

Hence, the aim of the current study is to delineate the IRS/CIRS and erythron profiles of chronic fatigue and affective symptoms due to MS and remitted RRMS patients.

## Material and Methods

### Participants

In the present case-control study, sixty-three patients with MS were recruited at the Neuroscience Center of Alsader Medical City in Al-Najaf province, Iraq, from September 2021 to March 2022. A senior neurologist diagnosed patients according to the McDonald criteria (Polman et al., 2011). Most MS patients (n=55) were in the remitted phase of RRMS and eight patients suffered from SPMS. We also recruited 30 healthy individuals, either staff, friends of staff or medical workers as a control group from the same catchment area as the patients. All the individuals were free of (lifetime and current) axis-1 neuropsychiatric disorders including a major depressive episode, schizophrenia, bipolar disorder, psycho-organic disorders, and substance use disorders (except nicotine dependence), medical disorders including chronic fatigue syndrome, diabetes mellitus, cardiovascular, thyroid, renal, liver, gastrointestinal, oncologic disorders, and (auto)immune, other neuroinflammatory, and neurodegenerative diseases, including psoriasis, COPD, inflammatory bowel disease, Parkinson’s and Alzheimer’s disease.

RRMS patients or their parents/legal guardians and controls gave written signed consent before inclusion in the study. The approval to conduct the current study was obtained from the institutional ethics board of the College of Medical Technology, The Islamic University of Najaf, Iraq (doc. no 11/2021). In this study, we followed Iraqi and foreign ethics and privacy rules based on the guidelines of the World Medical Association Declaration of Helsinki, The Belmont Report, CIOMS guidelines, and the International Conference on Harmonization of Good Clinical Practice; our IRB adheres to the International Guideline for Human Research Safety (ICH-GCP).

### Clinical Assessment

A senior psychiatrist evaluated sociodemographic, neuropsychiatric, and clinical data through a semi-structured interview. The EDSS (Kurtzke, John F., 1983) was used for clinical evaluation of disabilities, while the MSSS (Roxburgh et al., 2005) was used to evaluate the progression of disability over time reflecting severity of MS. Activities of daily living (ADL) were determined using the Arabic translated index of ADL (Katz et al., 1970). In addition, the Hamilton Depression Rating Scale (HAMD) (Hamilton, 1960), Hamilton Anxiety Rating Scale (HAMA) (Hamilton, 1959) and the Fibro-Fatigue scale (Zachrisson et al., 2002) were used to determine severity of depressive, anxiety and fibromyalgia-fatigue symptoms, respectively. We asked participants who took part in the study to rate their depression, anxiety and FF complaints over the past three months preceding the study. Based on our previous publications (Al-Hadrawi et al., 2022; Al-Hakeim et al., 2022b; Almulla et al., 2021), we computed several symptom subdomain scores as z unit-based composite scores using the z scores of different items, namely: a) pure depressive symptoms computed as the sum of depressed mood + feelings of guilt + suicidal ideation + loss of interest (HAMA) + sadness (FF), depressed mood (HAMA) + sadness + discouraged about the future + feeling a failure + dissatisfaction + feeling guilty + disappointed in oneself + critical of onseself + suicidal ideation + crying + loss of interest + difficulty with decisions + look unattractive + work inhibition (BDI); b) pure anxiety (pure HAMA) as the sum of anxious mood + tension + fears + anxiety behaviour at interview; c) pure physiosomatic symptoms as a z unit-based composite score based on the sum (z scores) of anxiety somatic + gastrointestinal + genitourinary + hypochondriasis somatic sensory + cardiovascular + gastrointestinal (GIS) + genitourinary + autonomic symptoms + respiratory symptoms (HAMA symptoms) + muscle pain + muscle tension + fatigue + autonomic + gastro-intestinal + headache + malaise (FF scale) + fatigue (BDI) + anxiety somatic + somatic gastro-intestinal + general somatic + genital symptoms + hypochondriasis (HAMD); e) fatigue as the sum of fatigue (FF) + fatigue (BDI); autonomic symptoms as autonomic symptoms (FF and HAMA) and sleep disorders as sleep disorders (FF) + early + middle + late insomnia (HAMD) + insomnia (HAMA) + sleep disturbance (BDI). We divided participant’s body weight in kilogram by the height in squared meter to obtain the body mass index (BMI).

### Biochemical assays

In the early morning (8:00-11:00 a.m.), 5 ml of venous blood sample was withdrawn from all participants by disposal syringe and divided into EDTA and serum tubes. A complete blood count (CBC) was performed on all subjects utilizing EDTA whole blood. Serum was obtained after centrifugation of the blood at 35000 rpm, then stored as small aliquots using Eppendorf tubes and frozen at −80 ℃ until thawed for biomarker assays. We used a haematology analyzer (HumaCount 30) provided by Human Company (Wiesbaden, Germany) to perform CBC. A Bio-plex Pro™ Human Chemokine Assays (Bio-Rad Laboratories, Inc. (Hercules, USA) was employed to analyze a panel of cytokines and chemokines, including IL-1β, IL-1ra, IL-2, IL-4, IL-5, IL-6, IL-7, IL-8, IL-9, IL-10, IL-12(p70), IL-13, IL-15, IL-17, Eotaxin, fibroblast growth factor (FGF) basic, granulocyte colony-stimulating factor (G-CSF), granulocyte-macrophage colony-stimulating factor (GM-CSF), IFN-γ, IP-10, MCP-1(MCAF), macrophage inflammatory protein-1 (MIP-1)-α, PDGF-bb, MIP-1β, regulated upon activation, normal T cell expressed and secreted (RANTES), TNF-α, and vascular endothelial growth factor (VEGFs). The procedure was as follows: a) using sample diluent (HB) to dilute the serums (1:4), b) 50 microliters of the diluted samples were mixed with 50 µl of microparticle cocktail (containing cytokine/chemokines per well of a 96-well plate provided by the manufacturer) and incubated for 1 h at room temperature while shaking at 850 rpm, c) plate was rinsed three times, then we added 50 µl of diluted Streptavidin-PE to each well, and we incubated it (with shaking at 850 rpm) 10 minutes at room temperature. d) 125 µl of assay buffer was added, and the wells were shaken at room temperature (850 rpm) for 30 seconds before being read using the Bio-Plex® 200 System (Bio-Rad Laboratories, Inc.). We utilized the immunofluorecense (IF) values of the cytokines/chemokines in the data analyses (Maes et al., 2022; Thisayakorn et al., 2022). Cytokines/chemokines which showed > 35% out of range concentration levels were not entered in the analysis, although the IF values were used to compute z unit-based composite scores (see below) (Maes et al., 2022; Thisayakorn et al., 2022). Consequently, we did not include IL-2, IL-5, IL-7, IL-15, GM-CSF, and VEGF in the regression analyses when using separate cytokine/chemokines. The intra-assay CV values for all analytes were < 11.0%. In the current study, the primary outcomes were various immune profiles, including the M1 macrophage profile computed as z IL-1β + z IL-6 + z TNF-α + z CXCL8 + z CCL3 + z sIL-RA; Th1 as z IL-2 + z IL-12 + z IFN (interferon)-γ; Th2: z IL-4 + z IL-9 + z IL-13; Th17: z IL-6 + z IL-17; T cell growth (all factors that promote T cell growth): z IL-4 + z IL-7 + z IL-9 + z IL-12 + z IL-15 + z GM-CSF (Maes and Carvalho, 2018; Maes et al., 2022). Neurotoxicity was conceptualized as a composite score comprising neurotoxic cytokines/chemokines: z IL-1β + z TNF-α + z IL-6 + z IL-2 + z IFN-γ + z IL-17 + z CCL11 + z CXCL10 + z CCL3 + z CCL5 + z CCL2 (Thisayakorn et al., 2022). Based on a recent paper (Al-Hakeim et al., 2022a) and the available cytokines we also computed a Th17-axis index as z IL-6 + z TNF-α + z IL-17. The Th1/Th2 ratio was computed as zTh1 – z Th2.

## Statistical analysis

We employed SPSS windows version 28 to perform all statistical analyses. The comparisons between category-based variables were accomplished by analysis of contingency tables (Chi-square tests). The differences between study’s group in terms of continuous variables were determined by analysis of variance (ANOVA). In addition, we utilized Quade Nonparametric Analysis of Covariance (ANCOVA) to examine differences in the non-normally distributed immune activation variables while covarying for CIRS values, to estimate the levels of immune activation above and beyond that of CIRS activation. Pairwise comparisons among group means were analyzed (at p<0.05) to define differences between three study groups. Furthermore, corrections of multiple comparisons were made utilizing the false discovery rate (FDR) p-value (Benjamini and Hochberg, 1995). Multivariate regression analyses were conducted to examine the best immunological and erythron profiles predictors of the disability and symptom subdomain scores while allowing for the effects of age, sex, and BMI. In addition, we also used a stepwise automated approach with p-values of 0.05 for model entry and 0.1 for model elimination. We estimated the model statistics (F, df, and p values), total variance explained (R^2^), and standardized beta coefficients with t statistics and exact p-values for each predictor. Furthermore, variance inflation factor (VIF) and tolerance were examined to check collinearity and multicollinearity issues. The White and modified Breusch-Pagan homoscedasticity tests were used to check for heteroskedasticity. To define the best cytokines and erythron profiles that predict disability and subdomain scores while allowing for the effects of confounders, we employed automatic linear modelling analyses with best subsets and overfit prevention criterion. To normalize distribution of the data, we used logarithmic or rank inverse-normal transformations. The significance was assessed using two-tailed tests with a p-value of 0.05. A priori calculation of the sample size using G*Power 3.1.9.4 showed that using an effect size of 0.2 at p=005 (two tailed) and power = 0.08, the estimated sample size should be at least 65.

## Results

### Sociodemographic, clinical and blood parameters of study’s groups

**Table 1** displays the sociodemographic characteristics, clinical symptoms and blood parameters of the MS patients and healthy controls. There were no differences in age, marital status, BMI, smoking between MS patients and controls. There were somewhat more females in the MS sample and more unemployed participants. This table also shows the measurements of MSSS, EDSS and ADL scores in patients and controls. The latter data show that our patients have mild disabilities and symptoms. We were able to extract one PC from the EDSS, MSSS and ADL scores (KMO=0.617, Bartlett’d test of sphericity χ^2^=126.03, df=3, p<0.001; AVE=73.57, all loadings >0.745). This PC reflects an integrated index of disabilities due to MS (labeled as PC_disabilities).

**Table 1:** Sociodemographic and blood parameters of multiple sclerosis (MS) patients and healthy controls (HC)

| **Variables** | **HC (n=30)** | **MS patients (n=63)** | **F/X<sup>2</sup>** | **df** | **p-value** |
| --- | --- | --- | --- | --- | --- |
| Age (Years) | 31.70(5.38) | 29.76(8.40) | 1.33 | 1/91 | 0.252 |
| Sex (M/F) | 19/11 | 23/40 | 5.90 | 1 | 0.015 |
| Marital status (Married/Single) | 19/11 | 29/34 | 2.43 | 1 | 0.119 |
| Employment (Y/N) | 28/2 | 11/52 | 48.04 | 1 | <0.001 |
| Smoking (Y/N) | 5/25 | 9/54 | 0.090 | 1 | 0.764 |
| BMI (Kg/m<sup>2</sup>) | 26.57(4.53) | 24.86(3.56) | 3.92 | 1/91 | 0.051 |
| Duration of illness (years) | - | 5.51(4.72) | - | - |  |
| MSSS | 0 | 2.08 (2.01) | MWU | - | <0.001 |
| EDSS | 0 | 1.06 (0.25) | MWU | - | <0.001 |
| ADL | 14.00 | 13.06 (1.40) | MWU | - | <0.001 |
| PC_disabilities | -0.967(0) | 0.518(0.659) | 151.41 | 1/91 | <0.001 |
| Relapsing remission / Progressive MS | - | 55/8 | - | - | - |
| WBC (thousand/mm<sup>3</sup>) | 6.59(1.75) | 9.49(2.96) | 24.59 | 1/91 | 0.009 |
| RBC (million/mm<sup>3</sup>)* | 4.915(0.111) | 4.926(0.086) | 0.00 | 1/81 | 0.940 |
| Hct (%)* | 42.974(0.947) | 40.838(0.732) | 3.08 | 1/81 | 0.083 |
| Hb (g/dl)* | 14.702(0.337) | 14.175(0.261) | 1.48 | 1/81 | 0.227 |
| MCV (fl)* | 87.931(1.249) | 84.144(0.965) | 5.58 | 1/81 | 0.021 |
| MCH (pg)* | 30.147(0.525) | 28.269(0.405) | 6.20 | 1/81 | 0.015 |
| RDW-SD (fl)* | 36.943(0.955) | 41.322(0.738) | 12.73 | 1/81 | <0.001 |
| PC_RBCs* | 0.189(0.153) | -0.041(0.118) | 1.37 | 1/81 | 0.245 |
| PC_RBCindices* | 0.332(0.183) | -0.084(0.141) | 3.15 | 1/81 | 0.080 |
| Pure depression | -0.964 (1.002) | 0.459 (0.592) | 73.62 | 1/91 | <0.0013** |
| Pure anxiety | -0.807 (0.645) | 0.384 (0.907) | 41.57 | 1/91 | <0.0013** |
| Pure physiosomatic | -1.223 (0.524) | 0.585 (0.524) | 243.61 | 1/91 | <0.0013** |
| Chronic fatigue | -0.912 (0.578) | 0.434 (0.856) | 60.68 | 1/91 | <0.0013** |
| Autonomic symptoms | -0.944 (0.518) | 0.449 (0.849) | 68.49 | 1/91 | <0.0013** |
| Insomnia | -0.824 (0.532) | 0.440 (0.860) | 63.54 | 1/91 | <0.0013** |
| PC_psychopathology | -1.228 (0.445) | 0.585 (0.561) | 240.69 | 1/91 | <0.0013** |

All results are shown as mean (SD); F/X<sup>2</sup>: results of analysis of variance and contingency tables, respectively; MWU: Mann-Whitney U (MWU) test; \* Results of factorial GLM analysis with sex as second factor (shown as mean and standard error); \*\*Results of false discovery rate p-correction
M: Male, F: Female, Y: Yes, N: No, BMI: Body Mass Index, Kg: Kilogram, m<sup>2</sup>: Squared meter, MSSS: Multiple sclerosis severity scores, EDSS: Expanded Disability Status Scale, ADL: Activities of daily living, Hb: Hemoglobin, Hct: Hematocrit, RBC: Red blood cells, MCV: Mean corpuscular volume, MCH: Mean corpuscular hemoglobin, RDW-SD: Red cell distribution width, WBC: White blood cells, dl: deciliter, g: gram, pg: picogram, fl: femtoliters, mm<sup>3</sup>: Cubic millimeter. PC\_disabilities: principal component extracted from ADL, MSSS and EDSS scores. PC\_RBCs: principal component extracted from RBCs, Hct and Hb. PC\_RBCindices: principal component extracted from MCV, MCH and RDW.

Since the erythron may be influenced by sex, we performed factorial GLM analyses with diagnosis and sex as factors. Age and BMI did not have any effect on these analyses, while RBC counts, Hb, Hct and PC_RBCs were significantly lower in females than in males (all p<0.001). We were able to extract one PC from the RBC, Hct and Hb data (KMO=0.684, Bartlett’d test of sphericity χ^2^=130.81, df=3, p<0.001; AVE=77.59, all loadings >0.856), which reflects cell counts, labeled PC_RBCs. We were able to extract one PC from the MCH, MCHC, and RDW-CV data (KMO=0.680, Bartlett’d test of sphericity χ^2^=89.045, df=3, p<0.001; AVE=73.62, all loadings >0.825), labeled as PC_RBCindices. Table 1 shows that there were no significant differences in RBCs, Hct, Hb and PC_RBCs between patients and controls. MCV and MCH were significantly lower in patients than in controls while RDW was significantly increased in patients, and there was a trend towards lowered PC_RBCindices. Since sex may influence the erythron parameters we checked consequent regression analyses by introducing sex as covariate.

All neuropsychiatric rating scale subdomain scores were significantly higher in patients than in controls. We were able to extract one latent vector from the subdomain scores (KMO=0.852, Bartlett’d test of sphericity χ^2^=278.00, df=15, p<0.001; AVE=62.24%, all loadings >0.678). This first PC extracted from the symptom domains is labeled PC_PP (PC_psychopathology). There was a significant association between PC_disabilities and PC_PP and all symptom domains scores (all r>0.515, p<0.001, n=93) in the combined group of patients and controls. In the total group combined (r=0.686, p<0.001, n=93) and in the restricted group of patients (r=0.339, p=0.006, n=63) there were significant associations between PC_disabilities and PC-PP.

### Assessment of immune profiles

Since we observed that a large part of the patients, but not all, showed increased IRS levels, we have performed clustering analyses to discover whether a valid cluster of patients with signs of IRS activation may be retrieved. Therefore, we conducted a two-step cluster analysis with patients versus healthy controls as the categorical variable and M1, Th1, Th2, Th17 and Treg phenotypes as clustering variables. With a good silhouette measure of cohesion and separation of 0.62, three clusters were generated. These groups are healthy control (n= 30), MS patients with high (n= 29) and low IRS (n= 34) activation scores. **Table 2** shows the different immune profiles of the patients and healthy controls and the results of Quade’s nonparametric ANCOVA with pairwise comparisons among group means. Apart from confounders we also used CIRS as a covariate to examine the activation of different IRS phenotypes above and beyond CIRS activity. The current study’s results indicate that patients with high MS have significantly increased activity of M1, Th1, Th17, T cell growth, and growth factors as compared with the low MS group and healthy controls. However, no significant differences were found between MS patients with low MS and healthy controls. As such, two valid clusters are formed, one with a comparable immune profile as detected in controls, and a cluster with increased IRS activation. In addition, patients with high IRS activity showed increased neurotoxicity scores. Increased Th2 and CIRS responses were established in the low MS but not in the high MS groups. There were no differences in Th1/Th2 ratio between patients and controls and there were no differences in any of the immune profiles between RRMS and SPMS.

**Table 2:** Cytokine profiles of healthy controls (HC) and patients with multiple sclerosis (MS) patients divided into with increased (HIGH) and normal (LOW) immune activation

| Variables | HC (n=30)<sup>A</sup> | LOW MS (n=34)<sup>B</sup> | HIGH MS (n=29)<sup>C</sup> | F | dfh/dfe | p-value |
| --- | --- | --- | --- | --- | --- | --- |
| M1 (z scores)* | -0.347(0.079)<sup>C</sup> | -0.210(0.077)<sup>C</sup> | 0.605(0.174)<sup>A,B</sup> | 19.43 | 2/90 | 0.002 |
| Th1 (z scores)* | -0.329(0.096)<sup>C</sup> | -0.159(0.130)<sup>C</sup> | 0.528(0.119)<sup>A,B</sup> | 12.42 | 2/90 | 0.002 |
| Th2 (z scores) | -0.164(0.050)<sup>B</sup> | 0.135(0.062)<sup>A</sup> | 0.010(0.099) | 5.72 | 2/90 | 0.018 |
| zTh2-zTh1 (z scores) | -0.172(0.127) | 0.175 (0.187) | -.027 (0.211) | 0.986 | 2/90 | 0.377 |
| Th17 (z scores)* | -0.304 (0.092)<sup>C</sup> | -0.062(0.099)<sup>C</sup> | 0.388(0.186)<sup>A,B</sup> | 4.54 | 2/90 | 0.002 |
| Th17-axis (z scores)* | -0.909(0.218)<sup>C</sup> | -0.226(0.262)<sup>C</sup> | 1.20(0.443)<sup>A,B</sup> | 8.21 | 2/90 | 0.002 |
| IRS (z scores)* | -0.308(0.079)<sup>C</sup> | -0.212(0.090)<sup>C</sup> | 0.567(0.143)<sup>A,B</sup> | 20.35 | 2/90 | 0.002 |
| Tcell growth (z scores)* | -0.147(0.070)<sup>C</sup> | -0.216(0.076)<sup>C</sup> | 0.406 (0.168)<sup>A,B</sup> | 9.08 | 2/90 | 0.002 |
| Growth factors (z scores)* | -0.333(0.068)<sup>C</sup> | -0.243(0.103)<sup>C</sup> | 0.629(0.190)<sup>A,B</sup> | 12.90 | 2/90 | 0.002 |
| Neurotoxicity (z scores) * | -0.330(0.086)<sup>C</sup> | -0.141(0.099)<sup>C</sup> | 0.507(0.136)<sup>A,B</sup> | 15.65 | 2/90 | 0.002 |
| CIRS (z scores) | -0.225(0.076)<sup>B</sup> | 0.210(0.103)<sup>A</sup> | -0.013(0.174) | 3.26 | 2/90 | 0.044 |

Results of Quade's nonparametric ANCOVA with age, sex, smoking, body mass index as covariates and \* with CIRS as additional covariate; dfh =degrees of freedom for the hypothesis, dfe =degrees of freedom for error; data are expressed as mean (SE), i.e. estimated marginal means; <sup>A,B,C</sup>: pairwise comparisons among group means. Shown are the false discovery rate corrected p values.
M1: Macrophage M1, Th: T-helper, IRS: Immune inflammatory response, CIRS: Compensatory immune-regulatory system.

Part of the patients were treated with IFN-β or betapherone (n=40) natalizumab (n=15) and other (fingolimod: n=4, hydrocortisone: n=2, methylprednisolone: n=2). Even without FDR p-correction, there were no significant effects of any of the drugs on the immune variables and no differences in immune profiles between these three treatment groups. Moreover, there was no significant association between the three medication groups and the cluster-analysis derived MS groups (χ2=4.20, df=2, p=0.134).

### Prediction of disabilities and severity of MS by immune biomarkers

Regression #1 and #2 were performed on all participants (**Table 3**) and show that a significant part of the variance (42.2%) in PC_disabilities could be explained by Th1, CIRS and Th17 axis. Nevertheless, within the selected group of MS patients no such correlations could be established. Regression #2 displays that T cell growth, CIRS, and Th1 could explain a significant amount of the variance (53.3%) in MSSS, with all predictors being positively associated. **Figure 1** shows the partial regression of the MSSS score on the Th1 profile. In the selected group of patients with MS, 17.8% of the variance in the MSSS score was predicted by Th1 and CIRS functions combined. In the combined study group, we found significant inverse correlations between PC_RBCs and PC_disabilities (r=-0.264, p=0.011, n=93) and PC_MSSS (r=-0.251, p=0.015, n=93). Nevertheless, these effects and PC_RBCindices were not significant in the multiple regression analyses shown in Table 3.

**Figure 1.**
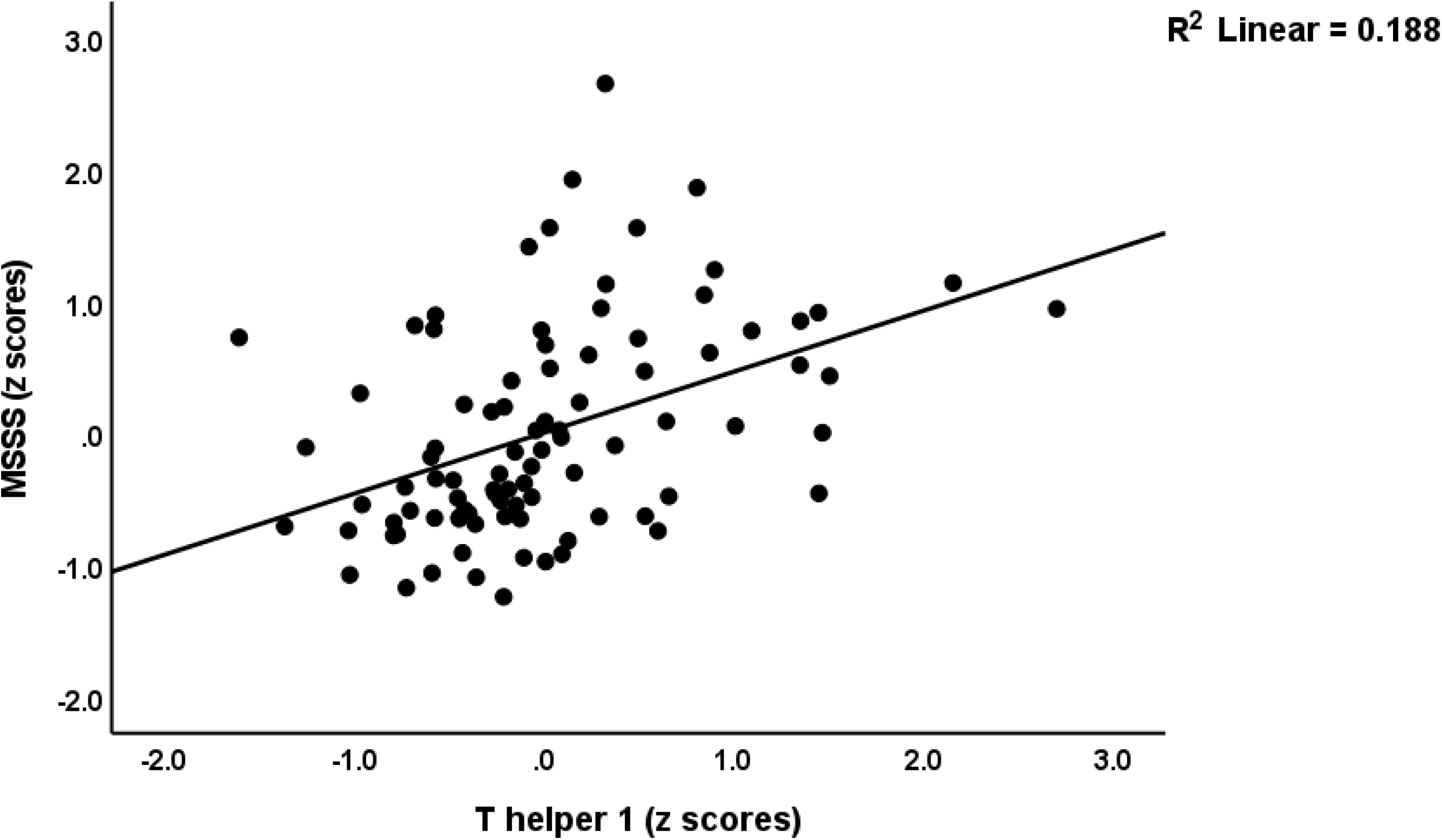
Partial regression of the multiple sclerosis severity scale (MSSS) scores on the T helper 1 profile.

**Table 3:**
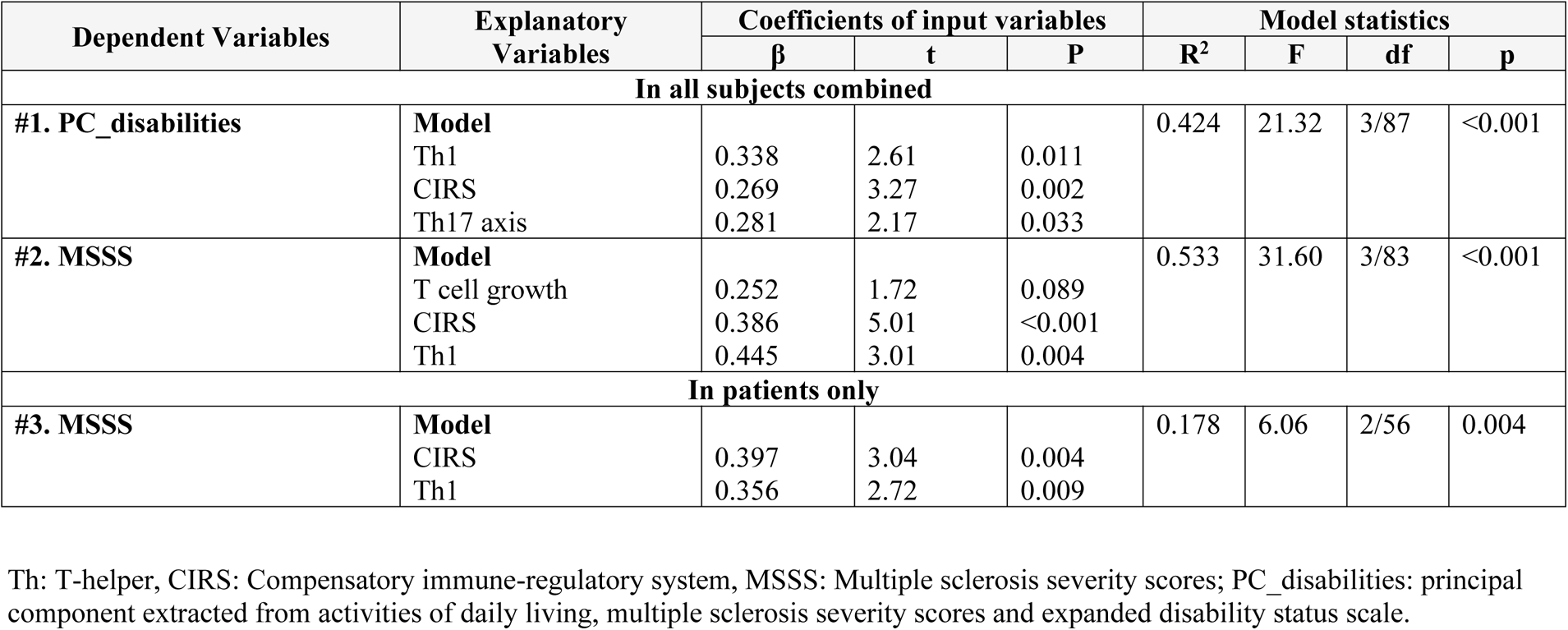
Results of multiple regression analyses with disabilities and multiple sclerosis severity scores (MSSS) as dependent variables and different immune profiles as explanatory variables.

| Dependent Variables | Explanatory Variables | Coefficients of input variables |  |  | Model statistics |  |  |  |
| --- | --- | --- | --- | --- | --- | --- | --- | --- |
| | | $\beta$ | t | P | R<sup>2</sup> | F | df | p |
| In all subjects combined |  |  |  |  |  |  |  |  |
| #1. PC_disabilities | Model |  |  |  | 0.424 | 21.32 | 3/87 | <0.001 |
|  | Th1 | 0.338 | 2.61 | 0.011 |  |  |  |  |
|  | CIRS | 0.269 | 3.27 | 0.002 |  |  |  |  |
|  | Th17 axis | 0.281 | 2.17 | 0.033 |  |  |  |  |
| #2. MSSS | Model |  |  |  | 0.533 | 31.60 | 3/83 | <0.001 |
|  | T cell growth | 0.252 | 1.72 | 0.089 |  |  |  |  |
|  | CIRS | 0.386 | 5.01 | <0.001 |  |  |  |  |
|  | Th1 | 0.445 | 3.01 | 0.004 |  |  |  |  |
| In patients only |  |  |  |  |  |  |  |  |
| #3. MSSS | Model |  |  |  | 0.178 | 6.06 | 2/56 | 0.004 |
|  | CIRS | 0.397 | 3.04 | 0.004 |  |  |  |  |
|  | Th1 | 0.356 | 2.72 | 0.009 |  |  |  |  |

Th: T-helper, CIRS: Compensatory immune-regulatory system, MSSS: Multiple sclerosis severity scores; PC\_disabilities: principal component extracted from activities of daily living, multiple sclerosis severity scores and expanded disability status scale.

### Prediction of neuropsychiatric (NP) symptoms by erythron variables and immune indices

We performed regression analysis with neuropsychiatric symptoms as the dependent variables and PC_RBCs, PC_RBCindices and immune profiles as explanatory variables (**Table 4**). Regression #1 shows that a large part of the variance (50.6%) in PC_PP was explained by Th17-axis, white blood cell counts (WBCs), CIRS (all positively associated) and PC_RBCs (inversely associated). Forced entry of sex in this regression analysis showed that sex was not significant (t=-0.52, p=0.607) and that PC_RBC remained significant (t=-2.55, p=0.013). **Figures 2 and 3** show the partial regression of the PC_PP on the Th17-axis and PC_RBCs, respectively. In addition, forced entry of MSSS in the analysis (regression #2) shows that 56.3% of the variance in PC_PP was explained by MSSS, WBCs and Tcell growth (positively associated) and sex. Regression #3 shows that when we included patients only, 17.1% of the variance in PC_PP could be explained by WBC (positive association) and PC_RBCs (inverse association). These associations remained significant after entering sex, which was not significant (t=0.52, p=0.603). Regressions #4 to #9 show the regression of the various subdomain scores on the cytokines and RBC profiles, while allowing for the effects of demographic data. The highest effect size was established for pure physiosomatic symptoms which showed that 50.3% of its variance could be explained by the Th17-axis, WBCs and CIRS (all positively associated) and sex (regression #4). PC_RBCs showed a significant effect on all subdomains except the pure physiosomatic, fatigue and sleep domains. These effects remained significant even after forced entry of sex in the regression analyses. A larger part of the variance in fatigue (41.7%) was explained by neurotoxicity, sex and CIRS. **Figure 4** shows the partial regression of fatigue on neurotoxicity.

**Table 4:**
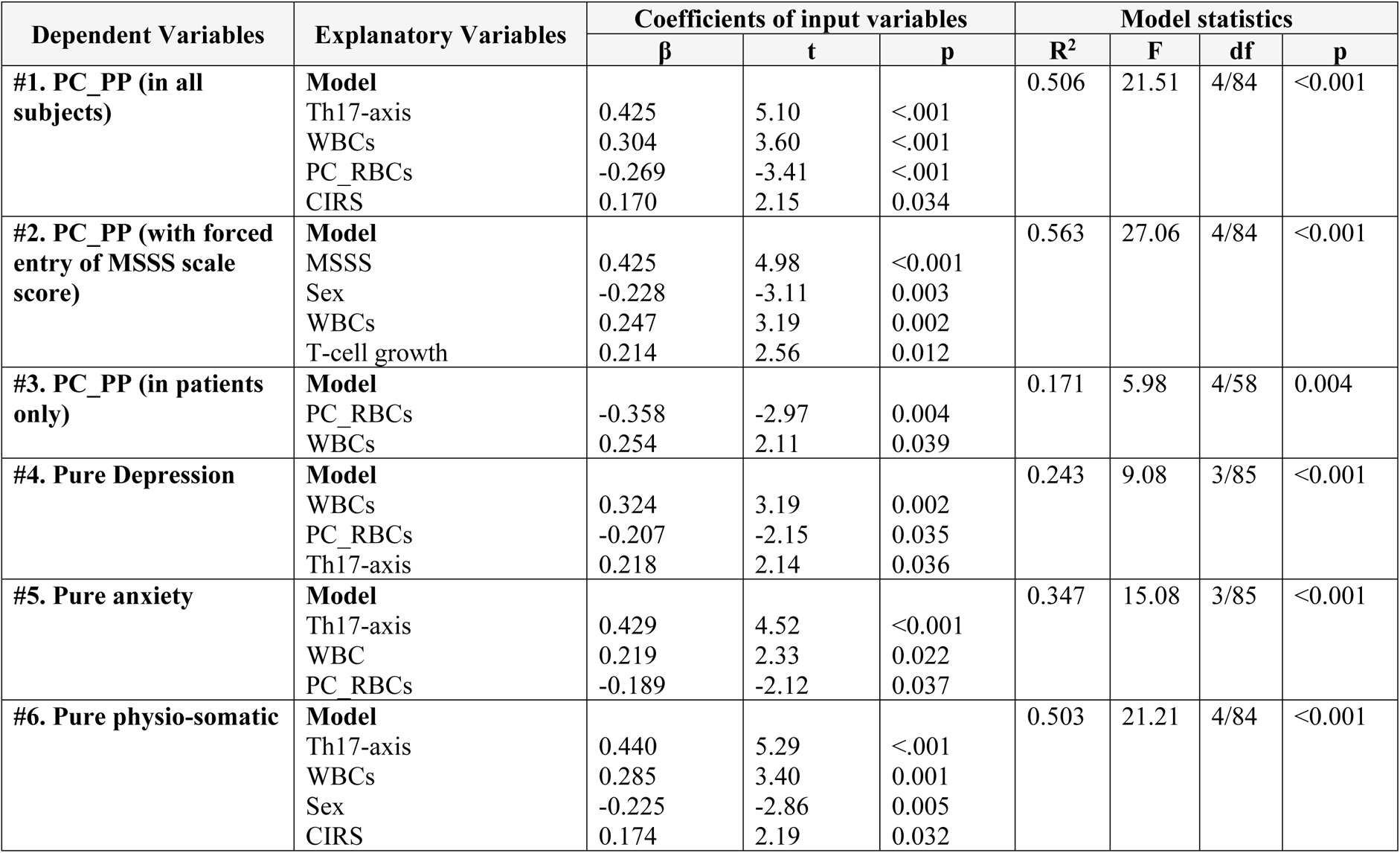
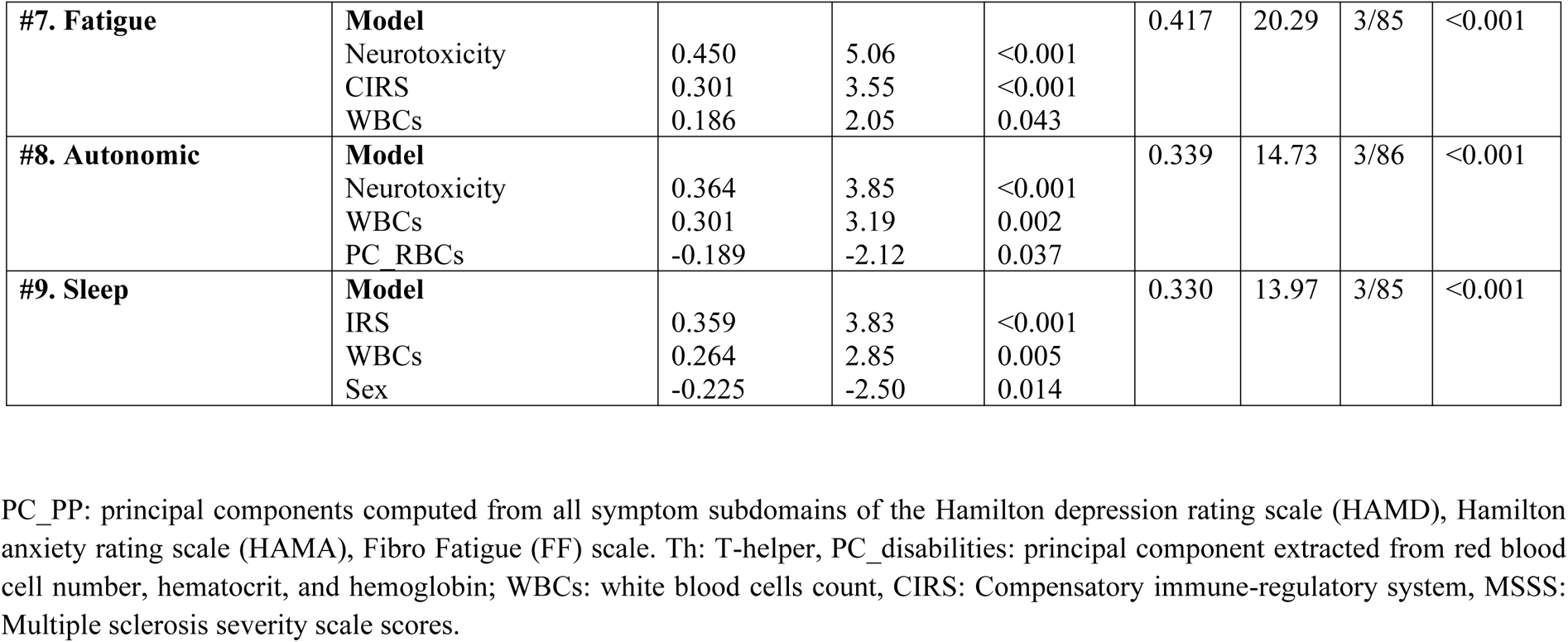
Results of multiple regression analyses with neuropsychiatric rating scale scores as dependent variables and immune profiles, erythron variables, and white blood cell (WBC) count as explanatory variables.

**Figure 2.**
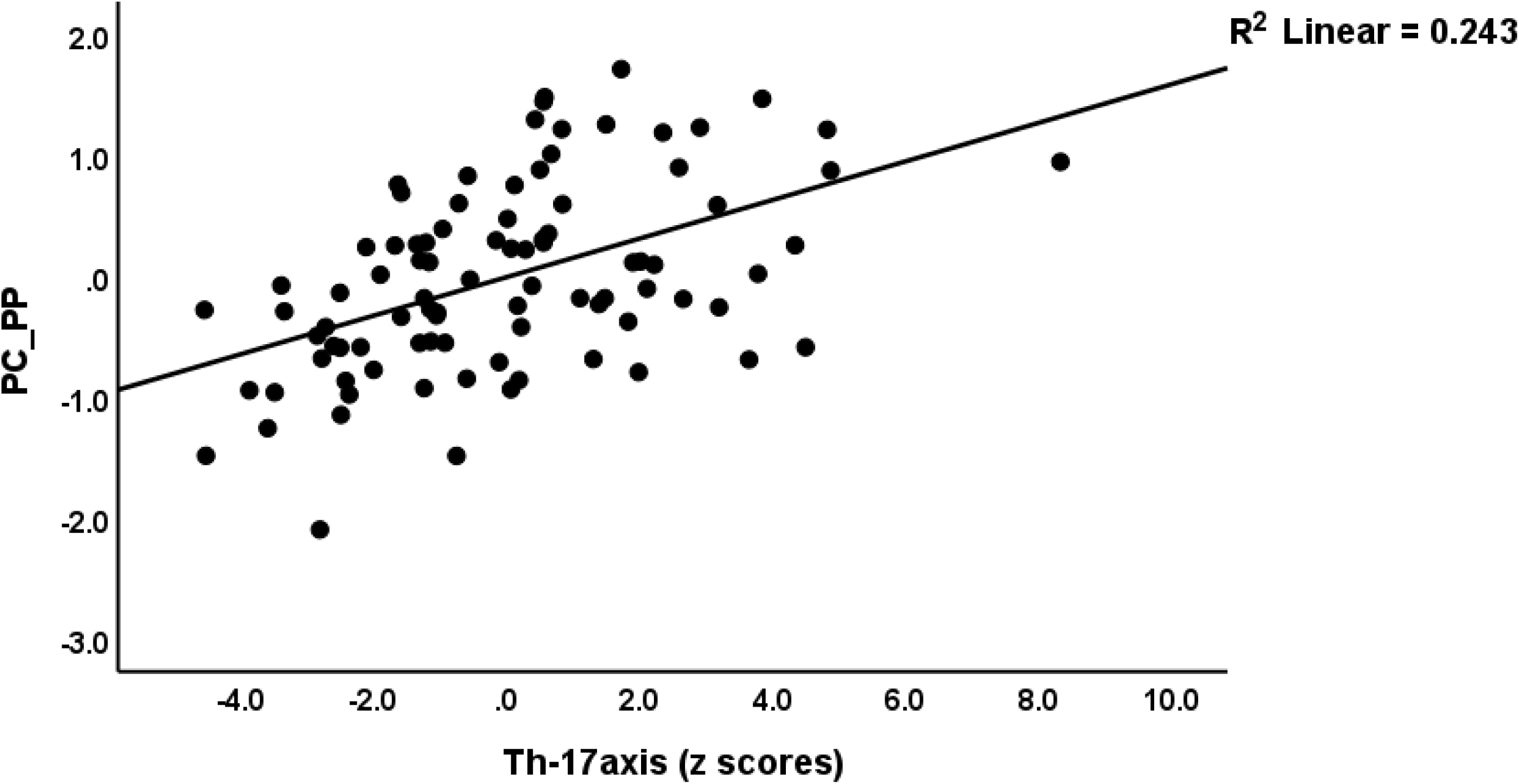
Partial regression of the first principal component score extracted from all psychopathology (PP) scores on the T helper 17-axis profile.

**Figure 3.**
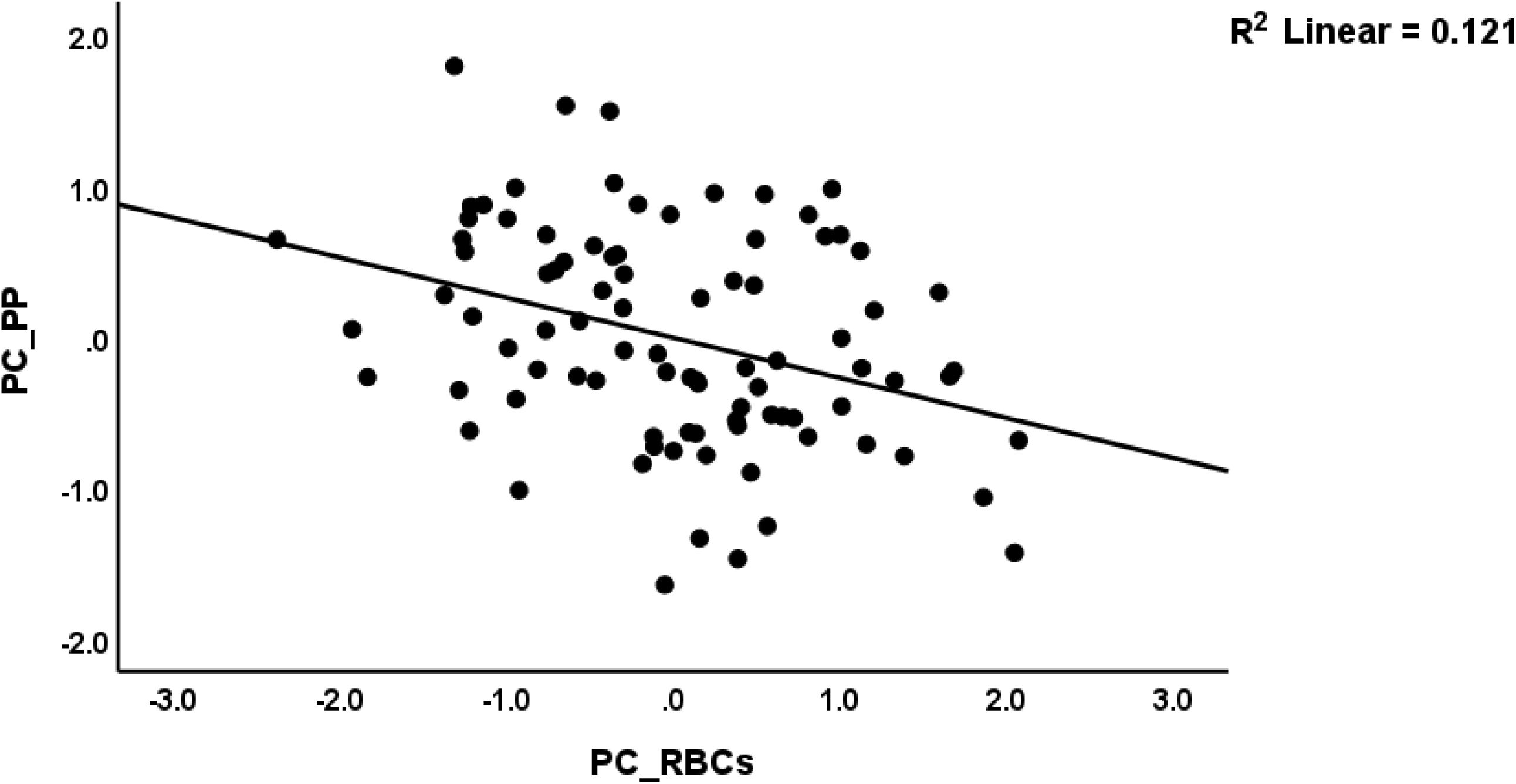
Partial regression of the first principal component score extracted from all psychopathology (PP) scores on the first PC extracted from number of red blood cells, hematocrit and hemoglobin (PC_RBCs).

**Figure 4.**
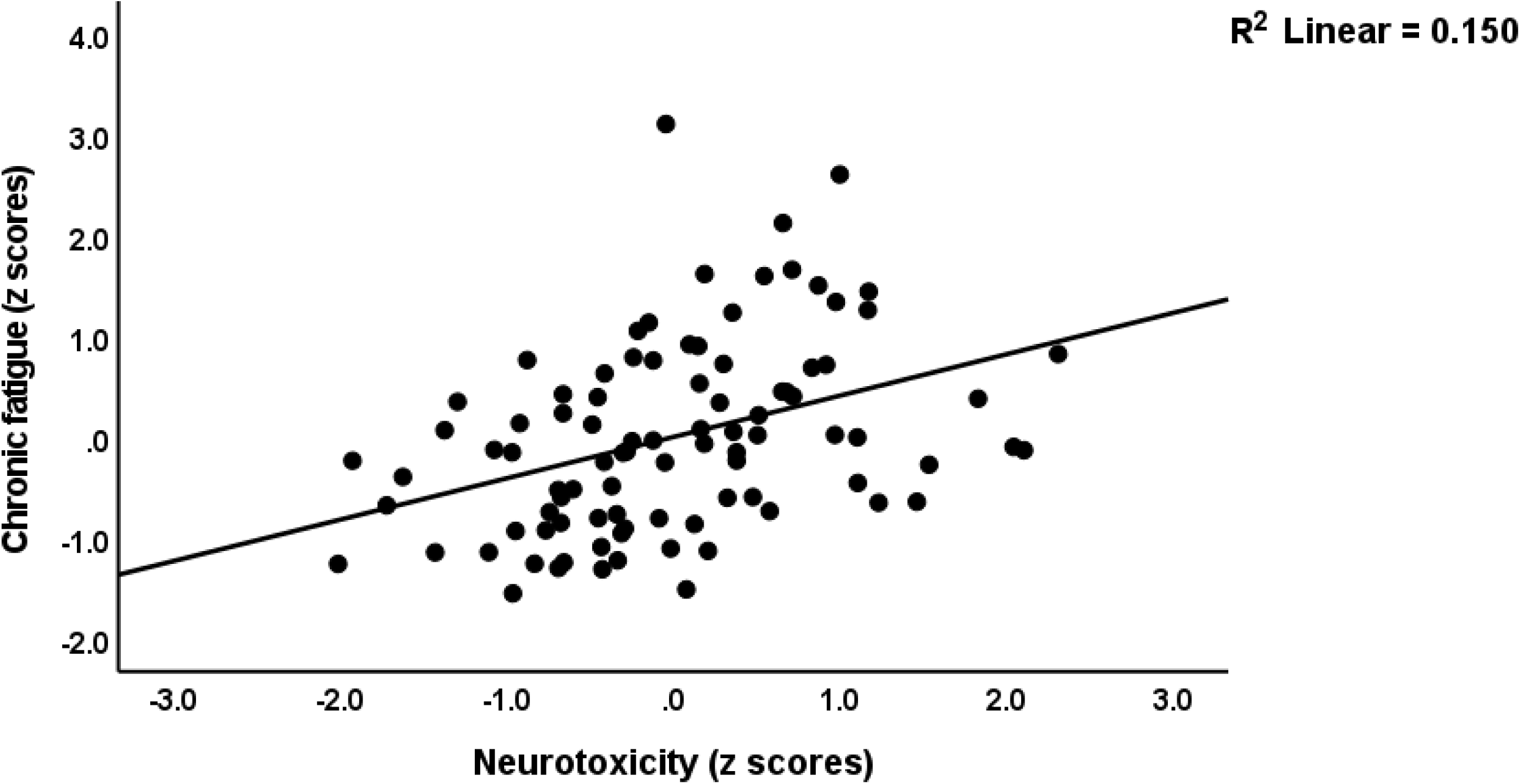
Partial regression of chronic fatigue symptoms due to multiple sclerosis on neuroimmune-toxicity.

### Results of automatic linear modelling analyses with overfit prevention

**Table 5** shows the results of automatic linear modelling, best subset with overfit prevention criterion, with PC_disabilities, PC_PP and the subdomain scores as dependent variables and the serum erythron variables, WBCs count, cytokines, chemokines and growth factors as explanatory variables while allowing for age, sex, BMI and smoking. Regression #1 shows that the best predictors of PC_disabilities are IFN-γ, IL-17 and sIL-RA (all positively associated), which together explain 45.9% of the variance in PC_disabilities. Regression #2 indicated that 47.4% of the variance in PC_PP score could be explained by WBC count, IL-10, and IL-6 (positively) and PC_RBCs (inversely). The best predictors of the subdomain scores were WBC count and different cytokines including IFN-γ, IL-4, IL-9, IL-10, IL-13, TNF-α, MIP1A, MCP1 (all positively), and PC_RBCs and sIL-1RA (both inversely).

**Table 5:**
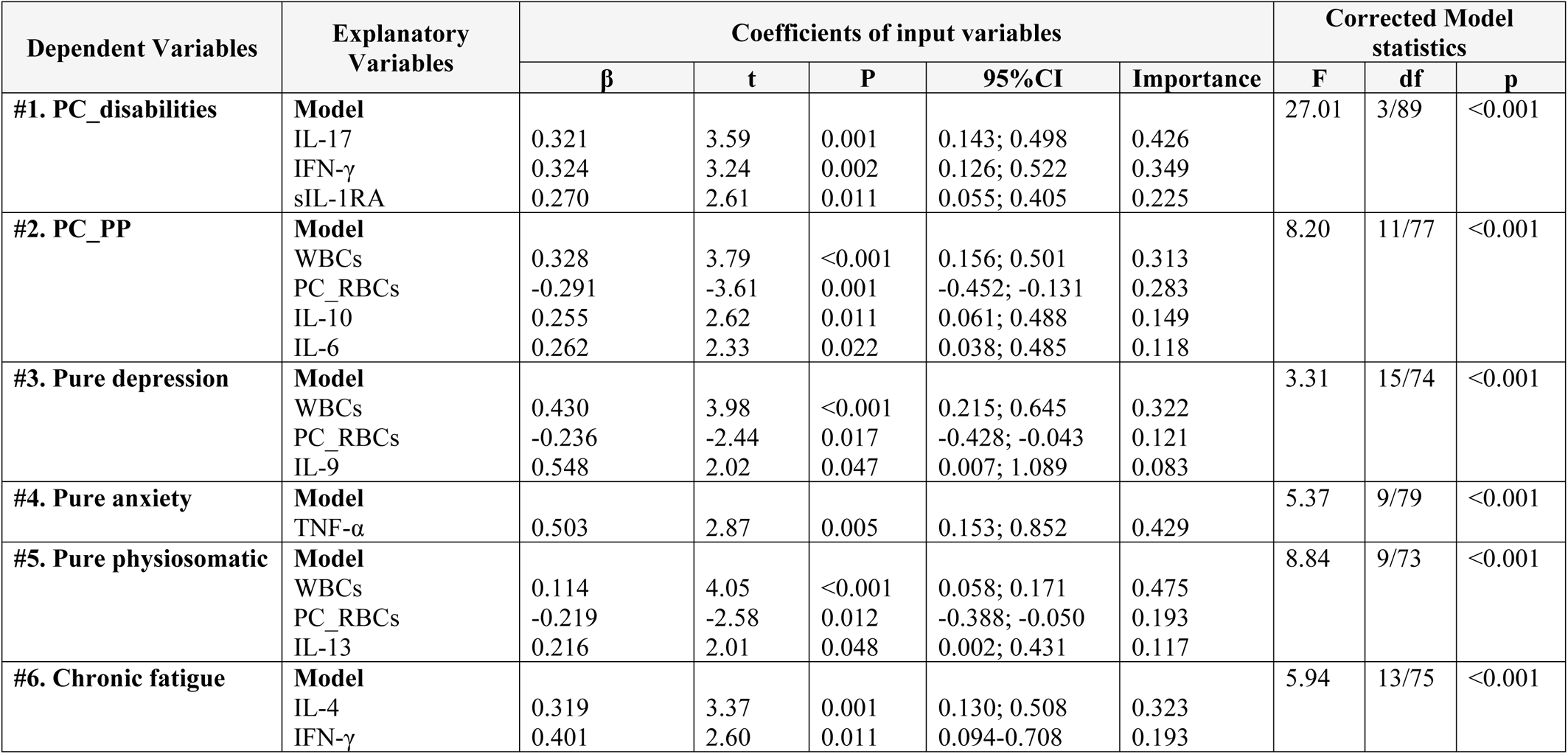
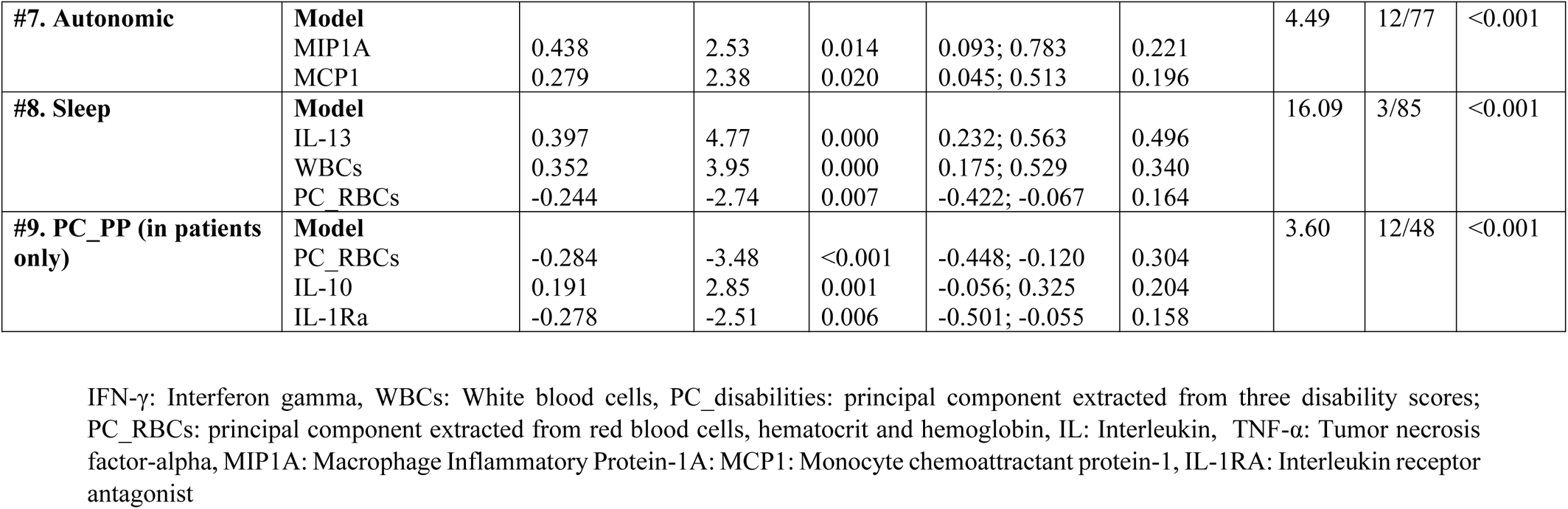
Results of automatic linear modelling analyses (best subsets with overfit prevention criterion) with multiple sclerosis disabilities and psychopathology (PP) rating scales and subdomain scores as dependent variables and serum erythron variables, white blood cell (WBC) count, cytokines, chemokines and growth factors as explanatory variables.

## Discussion

### Immune profiles in the remitted phase

The first major finding of the present research is that a significant part (50.9%) of remitted RRMS patients show activated IRS and CIRS, M1, Th1, Th17, T cell growth factor profiles. Moreover, we established that a combination of Th1 and Th17-axis activation with increased IFN-γ and IL-17 and increased CIRS indicators are associated with MS-related disabilities. As such, part of the remitted patients show an ongoing generalized immune-inflammatory process with increased neurotoxic capacity, indicating that the pathophysiological factors of RRMS are still active despite treatments with, for example, betapherone (IFN-1β) and natalizumab.

The current findings extend previous results reporting that significant elevations of central and peripheral Th-1/Th-2 cytokines, including TNF-α, IL-10, sIL-RA and other components of the IRS/CIRS are observed in RRMS patients of whom the majority are in the remission phase (Khaibullin et al., 2017; Rodríguez-Sáinz Mdel et al., 2002). Kallaur et al. detected a disbalance between Th1, Th2 and Th17 cytokines in MS-remitted patients (Kallaur et al., 2013). Hollifield et al., reported a significantly lowered biologically active TGF-β along with reduced IFN-γ, IL-1β, T-cell mitogen (PHA) and myelin basic protein in peripheral blood mononuclear cells (PBMC) of remitted MS patients (Hollifield et al., 2003). It is assumed that the active phase differs from the remission phase in terms of abnormal cytokine profiles (Morris, Gerwyn et al., 2018). For example, during the relapse phase of RRMS, TGF-β1 was significantly decreased while this cytokine was increased in the remission phase, indicating upregulated CIRS and Treg activities (Bertolotto et al., 1999; Mokhtarian et al., 1994; Rieckmann et al., 1995; Rieckmann et al., 1994). Moreover, Th17 is upregulated along with a significant increase of IFN-γ expressing Th17 lymphocytes in the CSF of MS patients during the relapse phase (Brucklacher-Waldert et al., 2009; Kebir et al., 2009). Furthermore, increased Th17 is frequently observed in RRMS (Kalra et al., 2020), and IL-17A and IL-17F are associated with the number of relapses (Babaloo et al., 2015).

The ongoing IRS/CIRS response with upregulated M1, Th1, Th17-axis profiles and IFN-γ production during the remitted phase may contribute to breakdown of the blood-brain barrier (BBB) and chronic neurotoxicity (Maes et al., 2021) and, thus, demyelination. CNS neuroinflammation may be aggravated by the migration of peripheral inflammatory mediators across the damaged brain endothelial cells of the BBB (Kamphuis et al., 2015). Moreover, immune activation, either IRS or CIRS, may cause viral reactivation which may play a role in MS. For example, increased CIRS activity may lead to immunosuppression which may reactivate latent viral infections. Following relapses, female but not male patients may display reactivation of Epstein-Barr virus (EBV) in B lymphocytes (Irizar et al., 2014). The higher relapse rate in RRMS is accompanied by increased expression of some human endogenous retroviruses (HERVs) (Morris et al., 2019). Reactivation of MS-associated retrovirus (MSRV) from the HERV-W family may drive IRS activation via Toll-Like Receptor activation although MSRV reactivation may also result from IRS activation (Morandi et al., 2015). It should be added that increased production of IFN-γ and IL-17, which is associated with increased disabilities during the remitted phase, may play a role in autoimmunity via different mechanisms (O’Shea et al., 2002).

Future research should examine whether the group of remitted patients who exhibit enhanced immune responses are those that will develop new relapses or will show an earlier relapse or progress into SPMS. If so, a more general activation of the immune system (activation of M1, Th1, IFN-γ, Th17 axis, IRS and CIRS profiles) should be the drug target to prevent new relapses rather than focusing on one aspect of the immune system.

### Immune profiles and depression, anxiety, and physio-somatic symptoms due to MS

The second major finding of the current study is that fatigue, depression and anxiety scores are significantly higher in remitted RRMS and SPMS patients as compared with healthy controls and that MS-related disabilities during the remitted phase are strongly associated with chronic fatigue, depression, anxiety, physiosomatic and autonomic symptoms, and insomnia. Since we assessed HAMD, HAMA and FF scales over the past three months prior to the study, the results indicate that small increases in disabilities are accompanied by increased chronic fatigue and chronic affective symptoms. During relapses, depressive symptoms are more prominent than in the remission phase (McCabe, 2005), although other reports show non-significant associations of depressive symptoms and different clinical phases (Jefferies, 2006).

Importantly, chronic fatigue and affective symptoms during the remission RRMS phase are largely predicted by the Th17-axis, increased CIRS and WBC numbers along with increased IL-10, IL-6, IL-9, IL-13, IL-4 and IFN-γ. As such, increased activation of the Th17-axis and CIRS during the remitted phase of RRMS may drive chronic fatigue and affective symptoms due to MS. Previous reports showed that IL-6 is positively associated with depressive symptoms due to RRMS (Frei et al., 1991; Kallaur et al., 2016; Koutsouraki et al., 2011) and PMS (de Carvalho Jennings Pereira, W. L. et al., 2020). Chronic fatigue syndrome is considered to be an immune-inflammatory disease with many pathophysiological similarities to multiple sclerosis, including a variety of immunological and neurological abnormalities, such as immune activation, immunosuppression, mitochondrial dysfunctions, elevated synthesis of nuclear factor-κB, autoimmunity as well as increased oxidative damage and decreased antioxidant capacity (Morris, G. and Maes, M., 2013). Likewise, depression is characterized by activated IRS, CIRS and oxidative stress pathways [see (Leonard and Maes, 2012; Maes and Carvalho, 2018; Maes, Michael et al., 2012; Moylan et al., 2013)].

Previously, it was discussed how increased IL-17, IL-6 and TNF-α (the components of the Th17-axis in the current study) contribute to major depressive disorder and chronic fatigue symptoms (Al-Hakeim et al., 2022a; Cui et al., 2021; Kim et al., 2021; Maes, M. et al., 2014; Maes and Carvalho, 2018; Melnikov, M. and Lopatina, A., 2022; Morris, G. and Maes, M., 2013; Morris, Gerwyn and Maes, Michael, 2013; Nadeem et al., 2017; Vieira et al., 2010). Interestingly, the first paper that reported increased TNF-α in major depression, found that the serum levels of this cytokine were higher in major depression than in MS (Mikova et al., 2001). Morris and Maes (2013) reviewed that, in MS, immune activation including activation of the Th17-axis components may drive chronic fatigue and physiosomatic symptoms.

The neurotoxic effects of the Th17-axis components, which play a key role in MS and fatigue and affective symptoms due to MS, comes from their role in peripheral inflammation, gut barrier and BBB breakdown, microglial activation, neuroinflammation, and neurotoxicity to CNS circuits (Al-Hakeim et al., 2022a; Maes, Michael et al., 2014; Maes et al., 2020; Morris, Gerwyn and Maes, Michael, 2013; Prajeeth et al., 2017). Activated immune-inflammatory pathways and consequent aberrations in brain structure and functions may render MS patients prone to develop mood symptoms and chronic fatigue syndrome (de Carvalho Jennings Pereira, W. L. et al., 2020; Feinstein et al., 2004; Morris, G. and Maes, M., 2013). It should be added that the increased fatigue and affective symptoms during acute relapses (Hanken et al., 2019; Khatibi et al., 2020) may be ascribed to activated immune-inflammatory and nitro-oxidative stress pathways (Maes, Michael et al., 2012; Maes, M. et al., 2012; Moore et al., 2012; Morris, Gerwyn and Maes, Michael, 2013; Ormstad, Heidi et al., 2020; Šabanagić-Hajrić et al., 2015).

### Fatigue and PP due to MS and the erythron

The third major finding of the present study is that chronic fatigue and affective symptoms due to MS, but not MS per se, are associated with deficits in the erythron, namely lowered number of RBCs, Hct and Hb, whereas alterations in RBC indices have less impact. On the other hand, aberrations in RBC indices (especially increased RDW) are a hallmark of MS, but not chronic fatigue and affective symptoms due to MS. Our findings extend previous results showing that the erythron profile of MS patients indicates altered RDW values (Groen et al., 2016) and that, in MS, there is a significant association between high RDW and EDSS scores (Peng et al., 2015). Other haemorheological features of MS comprise increased aggregation of RBCs due to increased peripheral inflammation (Groen et al., 2016). RBC numbers, Hb and Hct may differ significantly between RRMS and SPMS and are lower in RRMS compared to normal controls (Kasprzycka et al., 2019). Furthermore, major depression is associated with abnormal erythron parameters including decreased RBCs, Hct and Hb, probably as a consequence of the chronic mild inflammatory response during that illness (Maes, Michael et al., 1996; Vandoolaeghe et al., 1999). In chronic fatigue syndrome, RBCs are less deformable, and show lower membrane fluidity and zeta-surface charge as compared with RBCs of healthy controls (Saha et al., 2019). It is common knowledge that anemia and iron deficiency may cause chronic fatigue symptoms.

Our results suggest that aberrations in the erythron in MS may contribute to the pathophysiology of chronic fatigue and affective symptoms due to MS. First, disorders in the erythron resulting from activated IRS may induce the hypoxia-inducible factor (HIF) pathway leading to hypoxic damage (Deng et al., 2016) as repeatedly reported in fatigue, depression and anxiety (Zhao et al., 2017). Second, RBCs have important antioxidant defenses including superoxide dismutase, catalase and glutathione peroxidase activities (Jóźwik et al., 1997), whilst also hemoglobin may serve as an antioxidant (Jóźwik et al., 1997). Increased lipid peroxidation, which accompanies IRS activation (Maes et al., 2011), may suppress the antioxidant defenses in the RBCs and change the morphometric features of RBCs and functions of Hb, whilst these changes may be attenuated by administration of antioxidants (Revin et al., 2019). Moreover, the antioxidant capacity of RBCs is reduced in MS leading to increased oxidative stress (Groen et al., 2016), which play a key role in chronic fatigue syndrome and affective symptoms (Maes et al., 2011; Morris, G. and Maes, M., 2013). Thus, disturbances in RBCs and their antioxidant capacity coupled with high peripheral oxidative stress and changes in blood rheology may increase the vulnerability to oxidative damage and contribute to ischaemic tissue damage (Groen et al., 2016) and, therefore, to chronic fatigue and affective symptoms.

## Limitations

First, the results would have been more interesting if we had assessed biomarkers of oxidative stress including lipid peroxidation and RBC superoxide dismutase, autoimmune biomarkers, and neurotoxic tryptophan catabolites, which are induced during immune activation. Second, although this study was performed on a smaller study sample, the sample size was estimated a priori, based on a power of 0.8. Moreover, post-hoc computation of the achieved power, given the computed effect and sample sizes and alpha, show that the power of the regression analyses shown in table 4 ranges between 0.92 and 1.0. Third, future research should examine the differences in immune and erythron profiles between the remission and acute relapse phases of RRMS and examine whether RRMS patients with increased immune activation show a higher relapse rate or an earlier relapse as compared with without immune activation.

## Conclusions

A significant proportion of RRMS patients in remission exhibit active immune-inflammatory pathways and neuroimmune-toxicity. Activated immune-inflammatory responses and an aberrant erythron may explain the presence of chronic fatigue, affective symptoms, physiosomatic symptoms, insomnia, and autonomic symptoms throughout the remission period of RRMS. MS-related chronic fatigue and affective symptoms are caused by immunological activation and erythrocyte abnormalities. The more generalized IRS and CIRS activation, as well as the Th1 profile and Th17-axis, are therapeutic targets to prevent subsequent relapses in a subset of remitted RRMS patients. Erythron abnormalities are emerging therapeutic targets for the treatment of RRMS-related chronic fatigue, and physiosomatic and mood symptoms.

## Data Availability

After the authors have thoroughly utilized the dataset, the corresponding author (M.M.) will make all pertinent data available upon reasonable request.

## Acknowledgments

We would like to express our appreciation to the individuals who worked diligently to compile the data at the Neuroscience Center of Al-Sader Medical City of Najaf, Iraq.

## Ethical approval and consent to participate

The approval to conduct the current study was obtained from the institutional ethics board of the College of Medical Technology, The Islamic University of Najaf, Iraq (doc. no 11/2021). In this study, we also followed Iraqi and foreign ethics and privacy rules based on the guidelines of the World Medical Association Declaration of Helsinki, The Belmont Report, CIOMS guidelines, and the International Conference on Harmonization of Good Clinical Practice; our IRB adheres to the International Guideline for Human Research Safety (ICH-GCP). All participants provided written informed consent before enrollment in the study.

## Declaration of interest

The authors have no conflict of interest associated with the submitted article.

## Funding

The study was funded by the C2F program, Chulalongkorn University, Thailand. Grant Number: 64.310/169/2564

## Author’s contributions

AAA and AQA recruited the patients and performed the sampling. The measurements of serum biomarkers were performed by AFA. The study design and statistical analysis was carried out by MM. AFA and MM wrote and edited the manuscript. All authors contributed to the editing and approved the final version.

## Notes

### Competing Interest Statement

The authors have declared no competing interest.

### Author Declarations

The approval to conduct the current study was obtained from the institutional ethics board of the College of Medical Technology, The Islamic University of Najaf, Iraq (doc. no 11/2021).

